# Neutrophils Link Tissue Inflammation to Disease Control in Delayed Pressure Urticaria

**DOI:** 10.64898/2026.11.16.26368144

**Authors:** Marie Britz, Sangeetha Shankar, Franziska Wulfert, Meike Steinert, Caroline Constanze Albers, Michael Wojak, Soobin Dokko, Mathis Richter, Michael P. Schön, Mathias Sulk, Luise Erpenbeck

## Abstract

**Background:** Urticaria (hives) is a common inflammatory skin disorder affecting approximately 20% of the population during their lifetime. Chronic spontaneous urticaria (CSU) and delayed pressure urticaria (DPU) can substantially impair quality of life, and treatment remains challenging, particularly in refractory DPU. Although neutrophils are well-recognized components of urticarial skin infiltrates, their functional contribution and therapeutic relevance remain poorly understood.

**Objective:** To characterize neutrophil-associated inflammation in CSU and DPU, assess its relationship to disease control, and explore potential therapeutic targets.

**Methods:** Skin biopsies from patients with CSU (n = 4-17) and DPU (n = 5-15) and healthy controls (HC; n = 5) were examined by immunofluorescence for neutrophil infiltration, NET formation, and IL-17A and IL-23 positivity. Peripheral neutrophils were characterized by flow cytometry (CSU n = 8, DPU n = 5, HC n = 5) and circulating MPO-DNA complexes were measured by ELISA (CSU n = 33, DPU n = 29). Patients with refractory DPU received tildrakizumab on an individual off-label basis (n = 8). Disease activity, disease control and quality of life were assessed using the weekly Urticaria Activity Score (UAS7), the Urticaria Control Test (UCT) and the Chronic Urticaria Quality of Life Questionnaire (CU-Q2oL). Neutrophil activation markers were compared between patients receiving tildrakizumab (n = 8) and those without tildrakizumab treatment (n = 11). Paired measurements before and during treatment were evaluated in three patients.

**Results:** Despite similar overall dermal neutrophil infiltration, DPU showed greater neutrophil accumulation in the deep dermis than CSU. This spatial difference was accompanied by more pronounced NET formation in DPU. Higher NET-associated tissue signals correlated with poorer disease control in DPU, whereas no significant correlation was observed in CSU. Evidence of enhanced neutrophil activation in DPU also extended to the circulation, with increased CD66b and decreased CD62L expression compared with both CSU and HC. Circulating MPO-DNA levels were elevated in DPU relative to the HC reference, although the direct comparison between DPU and CSU was not significant. Investigation of the IL-23/IL-17 axis revealed IL-17A-positive neutrophils and IL-23-positive cells in lesional skin of both subtypes, with no significant difference in the proportion of IL-17A-positive neutrophils or IL-23-positive cells between CSU and DPU. In an exploratory therapeutic assessment, tildrakizumab treatment was associated with an overall significantly improved disease activity, control and quality of life in patients from week 0 to week 12. CD11b and CD66b expression was lower in treated than untreated patients, while paired analyses showed heterogeneous changes in neutrophil activation markers.

**Conclusion:** The combined tissue and blood findings suggest a more prominent role for neutrophil-associated inflammatory processes in DPU than in CSU. The presence of IL-17A-positive neutrophils and IL-23-positive cells in lesional skin provides a rationale for investigating the contribution of this cytokine pathway. Exploratory observations under tildrakizumab support further evaluation of IL-23 inhibition in refractory DPU, while its effects on neutrophil-associated inflammation require confirmation.

## INTRODUCTION

Urticaria, commonly known as hives, is a common inflammatory skin disorder affecting up to 20% of the population at least once in their lifetime (Haas et al., 1998; Lee et al., 2017; Zuberbier et al., 2022). It presents with transient pruritic wheals, angioedema, or both. Urticaria is classified as acute when symptoms occur for six weeks or less and as chronic (CU) when they persist or recur beyond this period (Lee et al., 2017; Zuberbier et al., 2022). CU comprises chronic spontaneous urticaria (CSU), in which symptoms arise without a definite eliciting stimulus, and chronic inducible urticaria (CIndU), in which symptoms are provoked by specific, reproducible triggers. A German population study reported CSU in 71.2% of patients with CU, CIndU in 19.7%, and both subtypes in 9.1% (Weller et al., 2022). Delayed pressure urticaria (DPU), a subtype of CIndU, is characterized by delayed swelling following sustained mechanical pressure (Zuberbier et al., 2022).

CU substantially impairs quality of life through recurrent wheals, angioedema, and pruritus, which contribute to psychological distress, sleep disturbances, and limitations in daily functioning (Gonçalo et al., 2021; Maurer et al., 2017). In DPU, pressure associated with routine activities such as walking, carrying bags, or prolonged sitting can trigger delayed swelling and discomfort, making avoidance of symptom-provoking stimuli difficult and further restricting daily activities (O’Donnell et al., 1997).

Non-sedating H1-antihistamines remain first-line therapy for CSU, with dose escalation recommended in antihistamine-refractory disease. Further treatment options, including the anti-IgE monoclonal antibody omalizumab, are available for patients who remain symptomatic despite updosing. Symptom control in DPU is considerably more difficult to achieve; responses to systemic corticosteroids or off-label dapsone are variable, and long-term corticosteroid use is limited by toxicity (Grundmann et al., 2011; Morioke et al., 2010). Notably, about one third of antihistamine-refractory patients with CU also fail to respond to omalizumab (Zuberbier et al., 2024), highlighting a substantial unmet clinical need. These therapeutic limitations are particularly relevant to DPU, whose underlying mechanisms remain poorly defined and for which data from controlled therapeutic trials are limited. A more detailed understanding of the inflammatory processes sustaining urticaria is therefore needed to guide the development of additional therapeutic targets.

Across urticaria subtypes, mast cell activation represents a common pathogenic denominator and is widely regarded as the central effector mechanism in CU. However, mast cells do not act in isolation, and accumulating evidence points to a contribution of other immune cell populations to disease pathogenesis. Neutrophils are a well-recognized component of urticarial infiltrates, yet their functional contribution remains poorly understood. Whether they simply accompany mast cell activation or actively amplify and sustain inflammation is therefore an important unresolved question, particularly in difficult-to-treat DPU.

Histopathologically, both CSU and DPU show mixed dermal inflammatory infiltrates comprising neutrophils, eosinophils, and lymphocytes, with neutrophils long recognized as a prominent component of urticarial lesions (Bernstein et al., 2014; Giménez-Arnau et al., 2021; Haas et al., 1998). Beyond their presence in the infiltrate, neutrophils can act as potent amplifiers of inflammation in skin disease (Neubert et al., 2019; Schön et al., 2017; Schön & Erpenbeck, 2018). One mechanism through which they exert these effects is the formation of neutrophil extracellular traps (NETs), extracellular chromatin structures decorated with antimicrobial proteins. Although NETs contribute to host defense by immobilizing and neutralizing invading pathogens (Fuchs et al., 2007), dysregulated NET formation has been implicated in inflammatory, autoimmune, and malignant diseases (Erpenbeck & Schön, 2017; Kahlenberg et al., 2013; Schön & Erpenbeck, 2018). NET formation must therefore be tightly regulated to prevent collateral tissue damage (Erpenbeck et al., 2019; Neubert et al., 2019).

These observations raise the question of whether neutrophils in urticarial lesions also undergo NET formation and whether this process is associated with disease control.

Neutrophils also interact directly with mast cells, providing a potential link between neutrophil-associated inflammation and the central effector mechanism of CU. Mast cell-derived TNF-ɑ promotes neutrophil activation and extravasation into inflamed skin (Dudeck et al., 2021). In addition, mast cells can internalize living neutrophils through the formation of mast cell intracellular traps (MITs) and subsequently release neutrophil-derived material, thereby prolonging inflammatory signal relay (Mihlan et al., 2024). These interactions provide a biological rationale for investigating how neutrophils contribute to the amplification and persistence of mast cell-driven inflammation in CU.

Beyond these cellular interactions, cytokine pathways that regulate neutrophil inflammatory responses offer potential therapeutic targets. The IL-23/IL-17 axis is well established in inflammatory skin disease, as demonstrated by the efficacy of therapies targeting this pathway in psoriasis (Griffiths et al., 2010; Langley et al., 2014). Neutrophils have been identified as an important source of IL-17A in inflammatory skin disease, particularly in association with NET formation (Keijsers et al., 2014; Lin et al., 2011). Experimental studies have further demonstrated that neutrophils can produce IL-17A under specific inflammatory conditions (Taylor et al., 2013; Li et al., 2018).

Clinical observations also implicate this pathway in urticaria. Elevated serum concentrations of IL-23 and IL-17 have been reported in patients with CSU, with both cytokines positively correlating with disease activity (Atwa et al., 2014; Sharma et al., 2020). IL-23 is of particular interest because it can regulate this neutrophil-associated cytokine response: together with IL-6, it induces IL-17A expression in human and murine neutrophils and supports an autocrine pathway that enhances neutrophil effector function (Taylor et al., 2013). Furthermore, IL-23 directly induces a cytokine-producing phenotype in murine neutrophils, characterized by IL-17A, IL-17F, and IL-22 expression (Li et al., 2018). Together, these findings raise the possibility that IL-23 contributes to urticarial inflammation by promoting inflammatory neutrophil responses. They provide a rationale for examining the relationship between IL-23 and neutrophil activation in CU and for exploring IL-23 inhibition as an upstream approach to modulating this inflammatory pathway.

Given the unmet clinical need, particularly in DPU, we sought to determine whether the established presence of neutrophils in urticarial lesions is accompanied by local and systemic activation and whether these features are associated with disease control. We therefore compared neutrophil distribution, NET formation, and peripheral activation profiles between CSU and DPU and examined IL-17A-positive neutrophils and IL-23-positive cells in lesional skin. Finally, we explored whether IL-23 inhibition with tildrakizumab was associated with clinical and biological changes in patients with therapy-refractory DPU.

## MATERIALS AND METHODS

### Patient Selection

Patients with confirmed CSU or DPU were enrolled, following ethical approval by the Landesärztekammer Westfalen-Lippe (reference 2022-420-f-S). DPU was confirmed by a positive pressure history and a positive standardized pressure test. Patients suspected of an alternative urticaria subtype (e.g., urticaria factitia) or with an ambiguous pressure test result were excluded. All patients were free from active infectious disease, severe autoimmune disease, or malignancy within the last 5 years at the time of sample collection. Peripheral blood was collected from urticaria patients attending the allergology outpatient clinic of the Department of Dermatology, University Hospital Münster. All patients had active urticaria with visible wheals at the time of blood collection and had discontinued urticaria medications three days prior to the appointment. Every patient completed the Urticaria Activity Score (UAS7), Urticaria Control Test (UCT) and the Chronic Urticaria Quality of Life Questionnaire (CU-Q2oL) to characterize disease activity and health-related quality of life.

### Ethics

Human skin and blood samples were obtained after written informed consent and with approval of the ethics committee of the Landesärztekammer Westfalen-Lippe (reference 2022-420-f-S), Department of Dermatology, University Hospital Münster, Germany. Participants retained the right to withdraw consent at any time without consequence.

### Skin tissue immunofluorescence

A 4-mm punch biopsy was obtained within 5 hours of lesion occurrence from lesional skin within an active wheal in symptomatic patients during active disease. Tissue samples were fixed in 4 % paraformaldehyde and embedded in paraffin according to standard protocols (Hollstein et al., 2023). Sections of 3 µm were cut and processed for immunofluorescence as previously described (Hollstein et al., 2023). Briefly, antigen retrieval was performed in EDTA buffer (pH 9; DCS, EL010C100) using a steamer for 20 minutes. The following primary antibodies were applied overnight at 4 °C: monoclonal anti-human myeloperoxidase (MPO; mouse IgA; 1:400; Proteintech 66177-1-lg) for neutrophils; monoclonal anti-human citrullinated histone H3 (H3cit; rabbit IgG; 1:250; Abcam ab5103) for NETosis; polyclonal anti-human IL-17A (goat IgG; 1:200; R&D Systems AF-317-NA); and monoclonal anti-human IL-23 (rabbit IgG; 1:200; Abcam AB314486). Secondary antibodies were applied for two hours at room temperature: anti-mouse AlexaFluor 647 (goat IgA; 1:500; Biozol/Southern Biotech SBA-1040-31), anti-rabbit AlexaFluor 488 (goat IgG; 1:500; Invitrogen A11034), anti-goat AlexaFluor 488 (donkey IgG; 1:500; Thermo Fisher A11055), and anti-rabbit AlexaFluor 488 (donkey IgG; 1:500; BioLegend 406416). Slides were mounted with DAKO Fluorescent Mounting Medium (DAKO S3023).

### Imaging and Quantification

The samples were images using Nikon Eclipse Ti-2 and Zeiss LSM microscope 980 and 800. The samples were subsequently analyzed using Fiji (version 2.16.0/1.54p; ImageJ, National Institutes of Health, Bethesda, MD). A Nikon NIS-Elements workflow was used to scan tissue sections and define regions of interest (ROIs) across dermal layers, guided by the upper and lower vascular plexus as anatomical landmarks. Exposure times and laser intensity and detector gain as applicable were kept constant across samples within each experiment. For Neutrophil infiltration and NETosis quantification, fluorescence signal area was quantified using a fixed color threshold applied uniformly to all images. Neutrophil infiltration was expressed as the MPO-positive area relative to the total measured area. NETosis rate was calculated as the H3cit-positive area relative to the MPO-positive area. All quantifications were performed in a blinded manner.

### Quantification of IL-17A–Positive Neutrophils and IL-23–Positive Cells

IL-17A–positive neutrophils and IL-23–positive cells were quantified by blinded manual counting of immunofluorescence images. For each dermal layer (superficial, mid, and deep dermis), three images were captured at 40,000× magnification with consistent exposure settings. Background correction was applied based on isotype control images.

IL-17A+ neutrophils were defined by MPO/IL-17A co-positivity and expressed relative to total MPO+ neutrophils. IL-23 positivity was determined relative to isotype controls. Three images per dermal layer and specimen were analyzed.

### Blood Collection and RBC lysis

Blood was drawn between 07:00 am and 12:00 pm to limit potential effects circadian variation in neutrophil activity and collected into 7.5 mL EDTA tubes Neutrophil isolation was initiated within 10 minutes of collection.

Red blood cell (RBC) lysis was performed on ice. Whole blood (200 µL) was incubated with 4 mL of 1× RBC lysis buffer on ice for 20 minutes. Lysis was terminated by dilution to 15 ml with EDTA supplemented Hank’s Balanced Salt Solution (HBSS). Cells were washed with PBS by centrifugation at 300 × g for 5 min at 4°C and resuspended in 150 µL HBSS.

### Flow Cytometry

Following RBC lysis, 5 × 10⁵ cells were Fc-blocked (BioLegend) for 10 min at 4°C and surface-stained with the indicated antibodies in Cell Staining Buffer (BioLegend) for 20 min at 4°C in the dark. The following antibodies were used: CD45 AlexaFluor 700 (BioLegend #304024), CD182 APC (#320710), CD184 APC/Cy7 (#306528), CD10 BUV395 (BD Biosciences #563871), CD16 BUV496 (#612944), CD97 BUV563 (#748366), CD15 BUV805 (BioLegend #315808), CD14 BV605 (#301834), CD62L PE (#304806), CD11a PE-Dazzle594 (#301231), CD49d PE-Cy5 (#304306), CD11b PerCP-Cy5.5 (#301328), CD66b Pacific Blue (#305112), and CXCR1 Spark Blue 550 (#285231).

Following surface staining, cells were washed, fixed and permeabilized using the Fixation/Permeabilization Kit (BioLegend) as per the manufacturer’s instructions. Intracellular staining was performed with IL-17A BV785 (BioLegend #512338), TNF-α BV711 (#502940), IL-8 PE-Cy7 (#511416), and IL-26 AF488 (R&D Systems #IC13751G) in Fix/Perm wash buffer. A total of 30,000 events were acquired per sample on a Cytek Aurora spectral flow cytometer. Data were unmixed using an established spectral unmixing matrix and analyzed with FlowJo v10 (BD Biosciences).

### MPO-DNA ELISA

Plasma was isolated by centrifugation at 1,500 × g for 10 minutes at 4 °C, aliquoted, and stored at -80 °C until analysis. Circulating NETs were quantified using a modified MPO-DNA ELISA optimized for human plasma. The assay captured NET complexes via an immobilized anti-MPO (mouse monoclonal clone 4A4, purified IgG; Bio-Rad, Cat. #0400-0002) antibody and detected NET-bound DNA using a peroxidase-conjugated anti-DNA antibody (Cell Death Detection ELISA PLUS, Roche Cat. No. 11 774 425 001). High-binding 96-well plates (Corning, 3590) were coated with 1 µg/ml of the capture antibody diluted in carbonate buffer overnight at 4 °C, blocked to reduce background with 4% BSA in PBS for 1.5 h at RT, and then incubated plasma samples overnight at 4 °C. After washing, plates were incubated with the anti-DNA-POD detection antibody for 1.5 h at RT, followed by TMB substrate (Thermo Fisher Scientific) development. The color development was stopped after 15 min with ELISA stop solution (Invitrogen, Thermo Fisher Scientific) and the absorbance was measured at 450 nm. As a reference, absorbance was also measured at 650 nm. All samples were run in duplicates and analyzed in batch to minimize interassay variability. This protocol enabled sensitive and reproducible detection of MPO-DNA complexes as a surrogate marker of NET formation.

The measured MPO-DNA signal was normalized to each patient’s daily peripheral neutrophil count (10³/µL) obtained from EDTA-anticoagulated whole blood and analyzed as part of routine clinical diagnostics at the University Hospital Münster.

For inter-assay normalization, pooled plasma from five healthy donors was used as an internal control. For statistical analysis, the pooled healthy control was defined as 100% baseline, and patient values were expressed relative to this reference.

### Statistical Analysis

Statistical analyses were performed using GraphPad Prism v7. Normality of data distribution was assessed using the Shapiro-Wilk test. Group comparisons were performed using Welch’s t-test, Mann–Whitney U test, or one-way ANOVA with multiple comparisons, as appropriate and specified in each figure legend. Correlations were assessed by Spearman rank correlation. Data are presented as mean ± SEM. A *P* value < .05 was considered statistically significant. Statistical significance is indicated as follows: *P* < .05 (*), *P* < .01 (**), *P* < .001 (***).

## RESULTS

### Neutrophil Infiltration and Spatial Distribution in CSU and DPU Skin

Although neutrophils are a recognized component of inflammatory infiltrates in both CSU and DPU, whether they exhibit distinct patterns of tissue involvement in these clinically different urticaria subtypes remains unclear. We therefore quantified the overall extent and spatial distribution of neutrophil infiltration in lesional skin biopsies. Overall neutrophil infiltration, assessed by the MPO-positive area fraction, was comparable between the two subtypes (Figure 1c). A pronounced difference emerged, however, when we examined the depth of infiltration: deep dermal neutrophil infiltration was significantly greater in DPU than in CSU (*P <* .0001; Figure 1d). These findings reveal distinct dermal neutrophil distribution patterns despite comparable overall infiltration, raising the possibility of subtype-specific differences in neutrophil recruitment or tissue localization.

**Figure 1:**
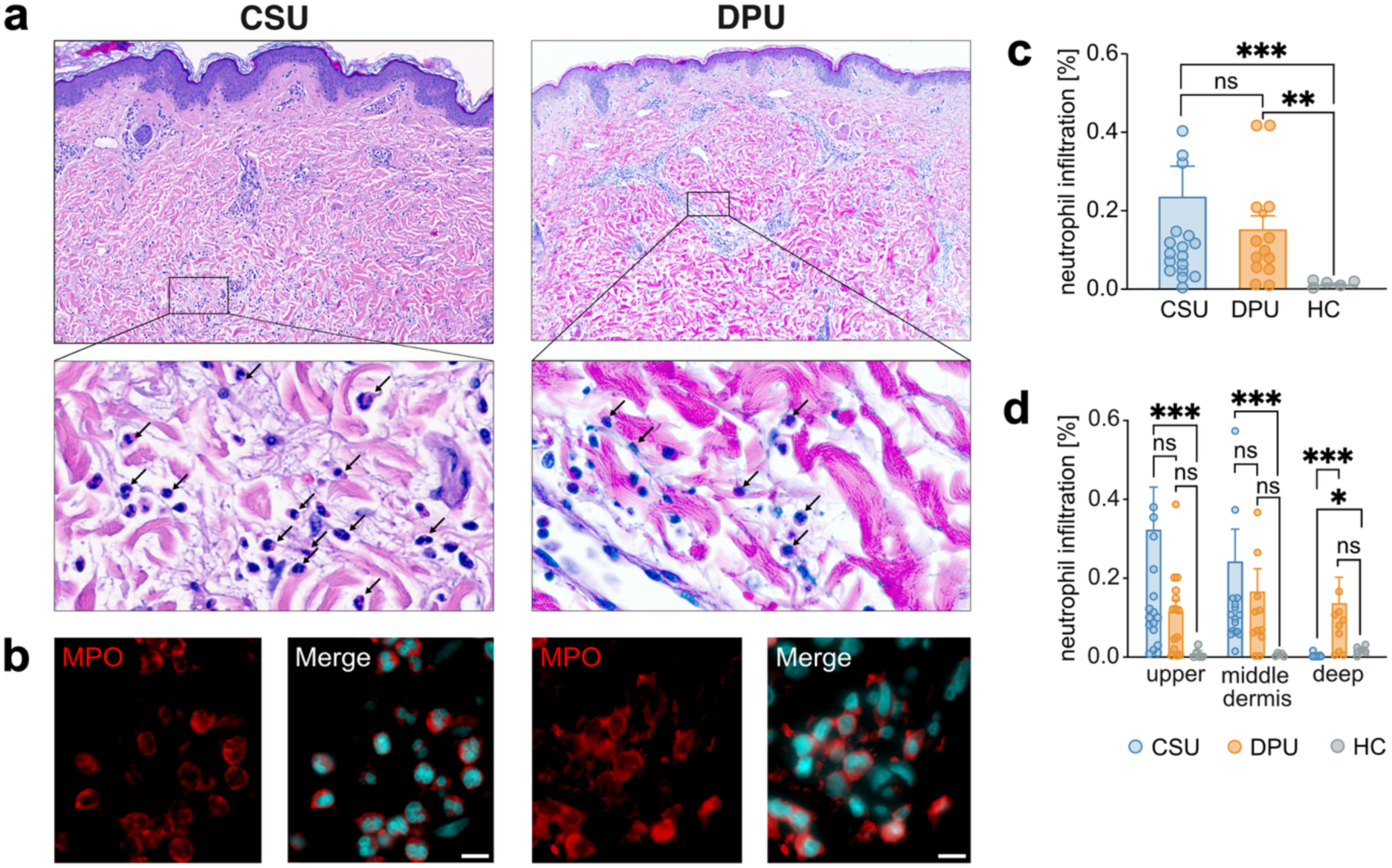
Neutrophil infiltration in CSU and DPU skin punches. **(a)** Representative hematoxylin and eosin (H&E)-stained sections of lesional skin from patients with DPU and CSU, demonstrating inflammatory infiltrates with prominent neutrophil accumulation (marked with black arrows). **(b)** Representative immunofluorescence images of lesional skin sections stained for myeloperoxidase (MPO, red) to identify neutrophils and DAPI (Chromatin, cyan) to visualize cell nuclei. Scale bar 1b = 10 µm. Neutrophil infiltration was quantified in immunofluorescence-stained skin sections by measuring MPO-positive area divided by total area and neutrophil infiltration is expressed as a percentage. **(c)** Neutrophil infiltration in total dermis (CSU n = 17; DPU n = 14; HC n = 5) **(d)** Neutrophil infiltration in upper, middle and deep dermis layer, as defined by anatomical structures. Data are presented as mean ± SEM from lesional skin biopsies of patients with chronic spontaneous urticaria (CSU; n [upper, middle, lower dermis] = 16, 16, 16) and delayed pressure urticaria (DPU; n [upper, middle, lower dermis] = 14, 11, 9), as well as from healthy control skin (HC, n = 5). Statistical analysis was performed using the Mann–Whitney U test. Data are presented as mean ± SEM.

### Lesional NET Formation Is Increased in DPU and Associated with Poorer Disease Control

These distinct infiltration patterns prompted us to ask whether neutrophils in CSU and DPU also differ in their local effector activity. We therefore examined NET formation in lesional skin and its association with disease control.

Immunofluorescence staining for MPO and citrullinated histone H3 (H3cit, a marker for NET formation) revealed NETs in lesional skin from both CSU and DPU patients (Figure 2a–b).

**Figure 2:**
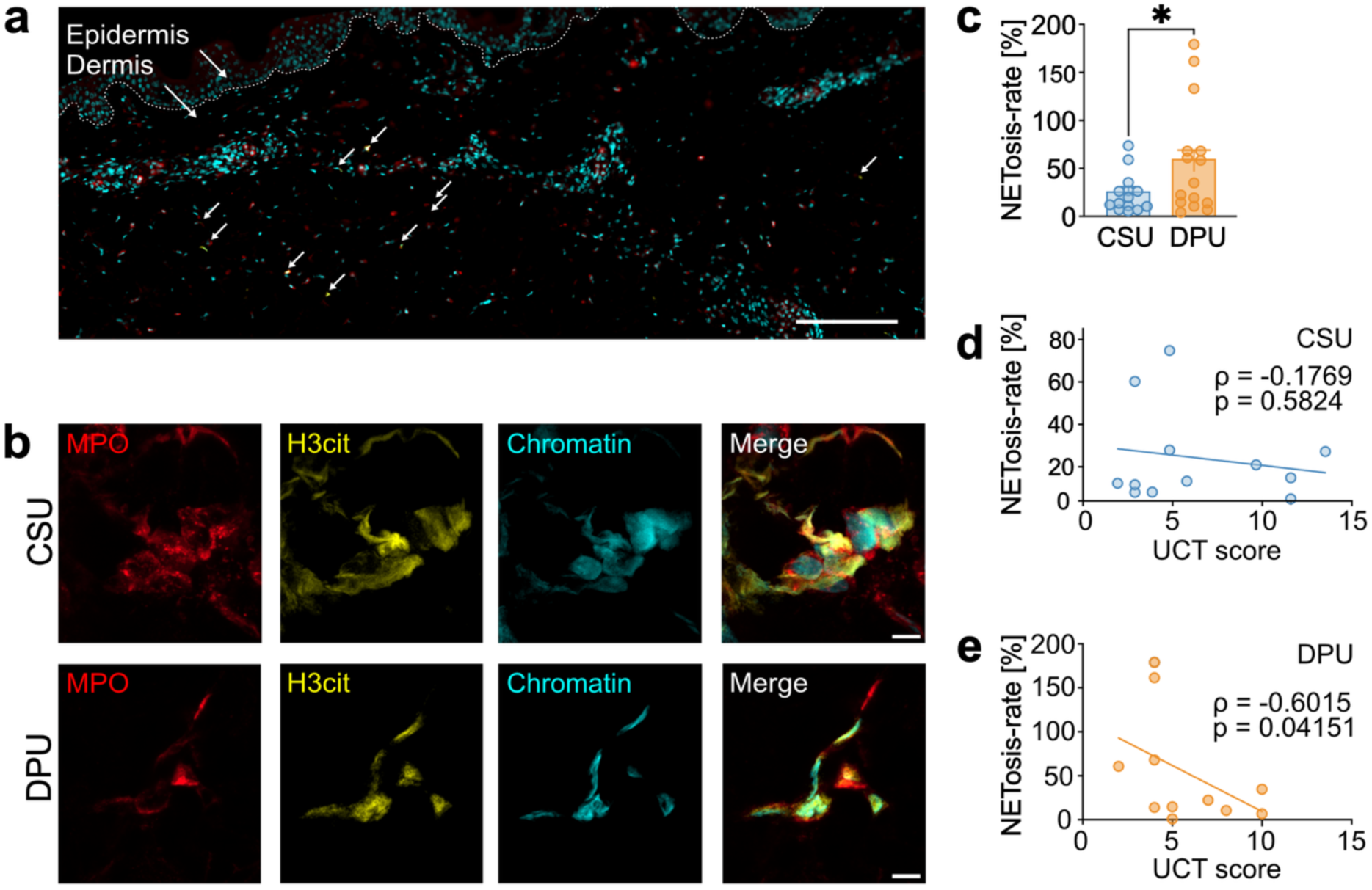
Images of immunofluorescence staining and quantification of neutrophil extracellular traps (NETs) in skin biopsies from CSU and DPU patients. (**a)** Overview image of the epidermis and dermis (separated by a dashed line) from a patient with CSU; representative neutrophil extracellular traps (NETs) are indicated by white arrows. **(b)** Confocal images of immunofluorescence staining in skin biopsies from CSU (top) and DPU patients (bottom). Fluorescently labeled antibodies against myeloperoxidase (MPO, red), H3cit (yellow), and DAPI (Chromatin, cyan), as a marker of NETosis. Scale bars: 3a = 100 µm; 3b = 5 µm. **(c)** NET formation in skin, as determined by evaluation of immunofluorescence in skin punches of CSU (n = 14) and DPU (n = 15) patients, evaluated as area^H3cit+^/area^MPO+^. **(d)** Correlation between NETs and disease activity measured by UCT score in CSU patients (n = 12) and **(e)** in DPU patients (n = 11). Spearman correlation analysis revealed a significant negative correlation (ρ = -.6015, *P* = .0415). Normality testing was performed prior to statistical analysis, followed by Welch’s t-test for group comparison. Data are presented as mean ± SEM.

Additional staining for neutrophil elastase (NE) confirmed the neutrophil origin of the extracellular chromatin structures (Supplementary Figure S1).

Despite comparable overall neutrophil infiltration, NET formation was significantly greater in DPU than in CSU (*P* = .047; Figure 2c), demonstrating a difference in local neutrophil effector activity between the two subtypes.

To assess the clinical relevance of this difference, we examined the association between lesional NET formation and disease control using the Urticaria Control Test (UCT). In DPU, NET formation correlated inversely with UCT scores (ρ = −.60, *P* = .042; Figure 2e), indicating that greater NET formation was associated with poorer disease control. No significant correlation was observed in CSU (ρ = −.18, *P* = .58; Figure 2d). Together, the greater extent of NET formation in DPU and its association with poorer disease control suggest that neutrophil effector activity may have a more prominent role in DPU than in CSU.

### Neutrophil Activation in Peripheral Blood

The increased lesional NET formation in DPU prompted us to investigate whether differences in neutrophil activation between CSU and DPU were also evident in the circulation. Using a customized flow cytometry panel, we assessed activation-associated surface markers on peripheral blood neutrophils from patients with active disease and healthy controls (HC). CD66b, a marker mobilized to the cell surface during specific granule exocytosis, was significantly more highly expressed in DPU than in both CSU and HC (DPU vs. CSU, *P* < .05; DPU vs. HC, *P* < .05; Figure 3a). Conversely, surface expression of the adhesion molecule L-selectin (CD62L), which undergoes proteolytic shedding upon neutrophil activation, was significantly lower in DPU than in either group (DPU vs. CSU, *P* < .01; DPU vs. HC, *P* < .01; Figure 3b). We also assessed CD11b, the α-subunit of the integrin Mac-1, which mediates adhesion and migration and is mobilized to the cell surface upon activation (Connelly et al., 2022). CD11b expression was higher in DPU, although neither comparison reached statistical significance (DPU vs. CSU, *P* = .532; DPU vs. HC, *P* = .064; Figure 3c). Together, these findings demonstrate that the differences between CSU and DPU extend beyond the distribution and effector activity of neutrophils in lesional skin to their activation phenotype in peripheral blood.

**Figure 3:**
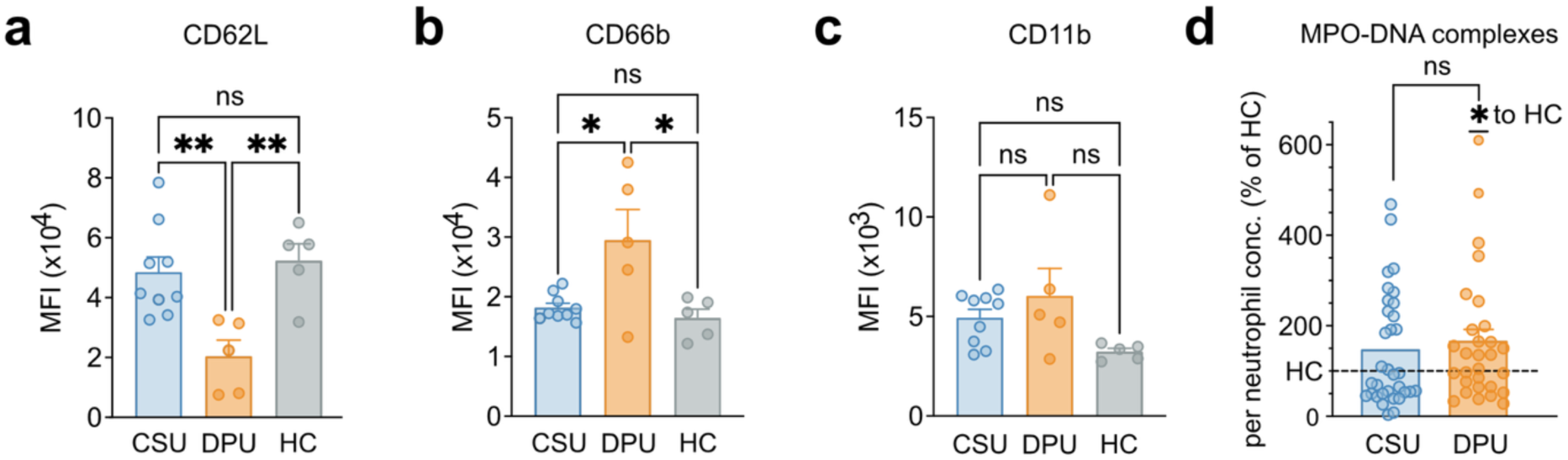
Neutrophil activation profiles in blood of CSU and DPU patients. Flow cytometry was performed on whole blood samples from patients with CSU (n = 9), DPU (n = 5) and HC (n = 5). Neutrophil activation was assessed by surface expression of (**a)** CD62L, **(b)** CD66b and **(c)** CD11b. Neutrophils were identified by sequential gating on CD45⁺ CD15⁺ CD16⁺ CD66b⁺ cells. Data are presented as mean fluorescence intensity (MFI). **(d)** NET formation in patients’ plasma was quantified by MPO–DNA complex measurements, normalized to the absolute peripheral blood neutrophil count at the time of blood sampling, and expressed relative to stimulated pooled healthy controls (n = 5), with HC defined as 100%, indicated by the dashed line. Normality was assessed prior to statistical analysis. Comparisons between CSU (n = 33) and DPU (n = 29) were performed using Welch’s *t*-test or the Mann–Whitney test, as appropriate. Comparisons with the HC reference (100%) were performed using a one-sample *t*-test or one-sample Wilcoxon signed-rank test, revealing significantly increased NET formation in DPU patients relative to the HC reference. Data are presented as mean ± SEM.

We next examined whether this circulating activation phenotype was accompanied by increased levels of circulating NET markers in plasma. MPO–DNA complexes, which are commonly used as a surrogate marker for NET-formation, were significantly elevated in DPU relative to the normalized HC reference (*P* = .042; Figure 3d), whereas in CSU NET markers did not differ significantly from HC (*P* = .181). Plasma MPO–DNA levels did not differ significantly between CSU and DPU (*P* = .477). Nevertheless, the combined findings of enhanced lesional NET formation, an activated circulating neutrophil phenotype, and elevated plasma MPO–DNA levels relative to HC support a more prominent contribution of neutrophils to inflammation in DPU.

### IL-23 and IL-17A Characterize the Pro-Inflammatory Milieu of Lesional Chronic Urticaria Skin

Having established that the neutrophil inflammatory phenotype in DPU extends from lesional skin to the circulation, we next sought to identify candidate signals that might sustain neutrophil-associated inflammation within the lesions. We therefore examined the local cytokine milieu, focusing on the IL-23/IL-17 axis. This choice was motivated by reports of elevated circulating IL-23 and IL-17 concentrations in CSU and experimental evidence implicating IL-23 in the regulation of inflammatory neutrophil responses, as outlined above.

Immunofluorescence staining identified IL-17A⁺MPO⁺ neutrophils in both CSU and DPU lesions (Figure 4a). No IL-17A-positive neutrophils were detected in HC skin, consistent with the absence of a neutrophilic infiltrate (Figure 1c). Among patients, the proportion of neutrophils positive for IL-17A did not differ significantly between CSU and DPU (*P* = .136; Figure 4b), indicating that neutrophil-associated IL-17A is a shared feature despite the differences in NET formation and peripheral activation described above.

**Figure 4:**
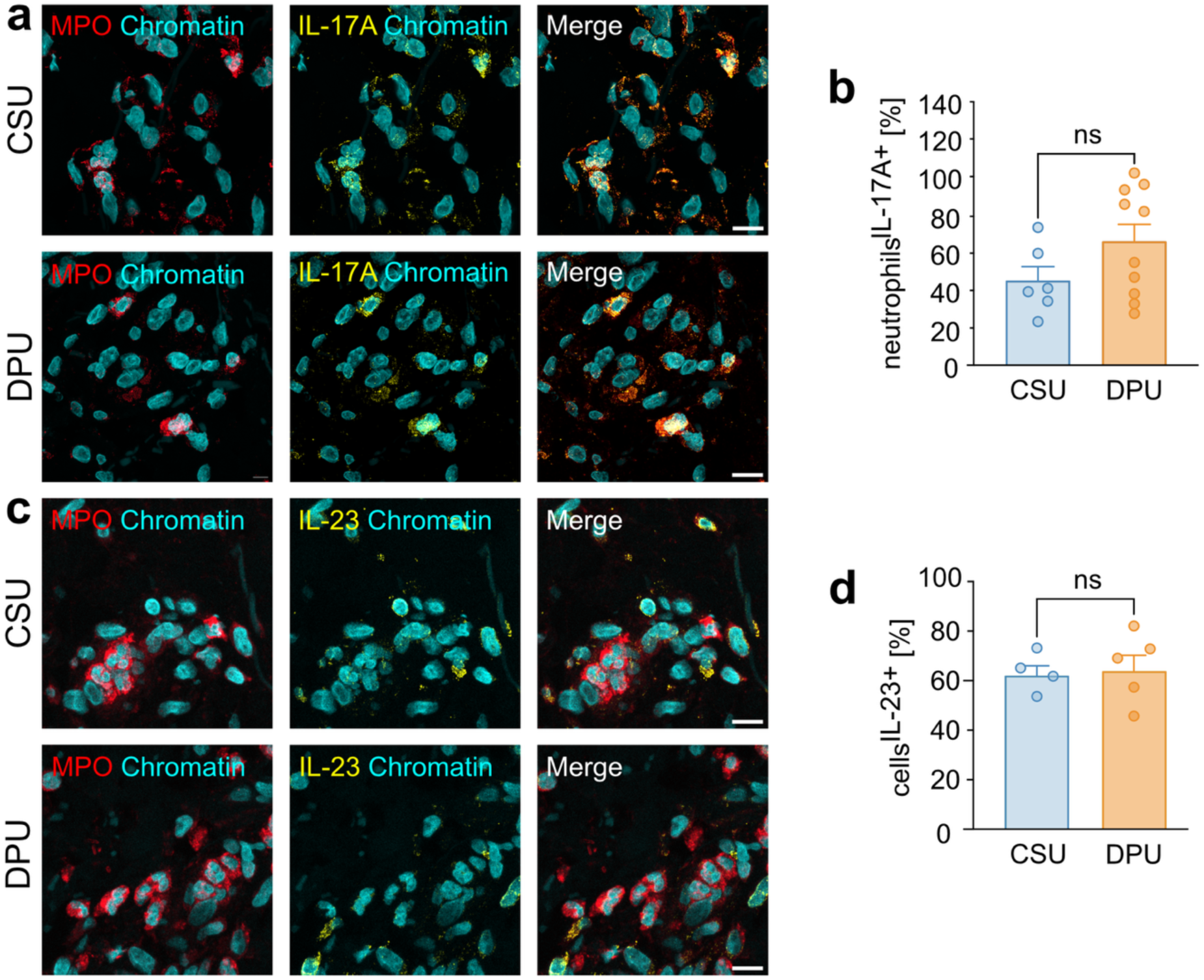
Immunofluorescence staining and quantification of IL-17A⁺⁺ neutrophils and IL-23⁺⁺ cells in skin biopsies from CSU and DPU patients. **(a)** Representative immunofluorescence images showing IL-17A⁺ neutrophils (MPO, red; IL-17A, yellow; DAPI-stained chromatin, cyan) **(b)** Quantification of IL-17A⁺ neutrophils expressed as the percentage of total MPO⁺ neutrophils in skin biopsies from patients with CSU (n = 6) and DPU (n = 5). **(c)** Representative immunofluorescence images showing IL-23⁺ cells (IL-23, yellow) in close proximity to MPO⁺ neutrophils (red); chromatin is stained with DAPI (Chromatin, cyan). Scale bars = 10 µm. **(d)** Quantification of IL-23⁺ cells as the percentage of total DAPI⁺ cells in skin from patients with CSU (n = 4) and DPU (n = 5). Scale bars 4a, 4c = 10 µm. Normality was assessed prior to statistical analysis. Group comparisons were performed using Welch’s *t*-test. Data are presented as mean ± SEM.

IL-23-positive cells were also detected in lesional skin, in close spatial proximity to MPO-positive neutrophils (Figure 4c). Additional staining identified monocyte/macrophage and dendritic cell populations among the IL-23-positive cells (Supplementary Figure S2-S3). Quantification of total IL-23-positive cells as percentage of all cells in CSU (n = 4) and DPU (n = 5) patients skin samples showed comparable IL-23-positivity rate (*P* = .9) (Figure 4c-d).

Together, the presence of IL-17A-positive neutrophils and their spatial proximity to IL-23-positive myeloid cells support the possibility of local paracrine interactions involving the IL-23/IL-17 axis in urticarial skin.

### Clinical and Biological Response to IL-23 Inhibition in Therapy-Refractory DPU

The presence of IL-17A-positive neutrophils and IL-23-positive myeloid cells in lesional skin, together with the pronounced neutrophil inflammatory phenotype in DPU, provided a rationale for exploring therapeutic intervention in this pathway. We selected IL-23 inhibition to target a potential upstream regulator of inflammatory neutrophil responses, including the acquisition of an IL-17-producing phenotype, rather than neutralizing IL-17A alone. This approach allowed us to explore whether targeting this regulatory pathway was associated with changes in both clinical disease control and neutrophil activation. We therefore assessed clinical outcomes and neutrophil activation in patients with therapy-refractory DPU receiving tildrakizumab on an individual off-label basis.

Clinical outcomes were assessed at baseline and weeks 4, 8, and 12. Tildrakizumab treatment was associated with significant improvements in disease activity, disease control, and quality of life. Between baseline and week 12, UAS7 scores decreased significantly (*P* = .02; Figure 5a) (n = 7; data from one patient were not evaluable due to incomplete diary entries), accompanied by increased UCT scores (*P* = .04; Figure 5b; n = 8) and decreased CU-Q2oL scores (*P* = .01; Figure 5c; n = 8).

**Figure 5:**
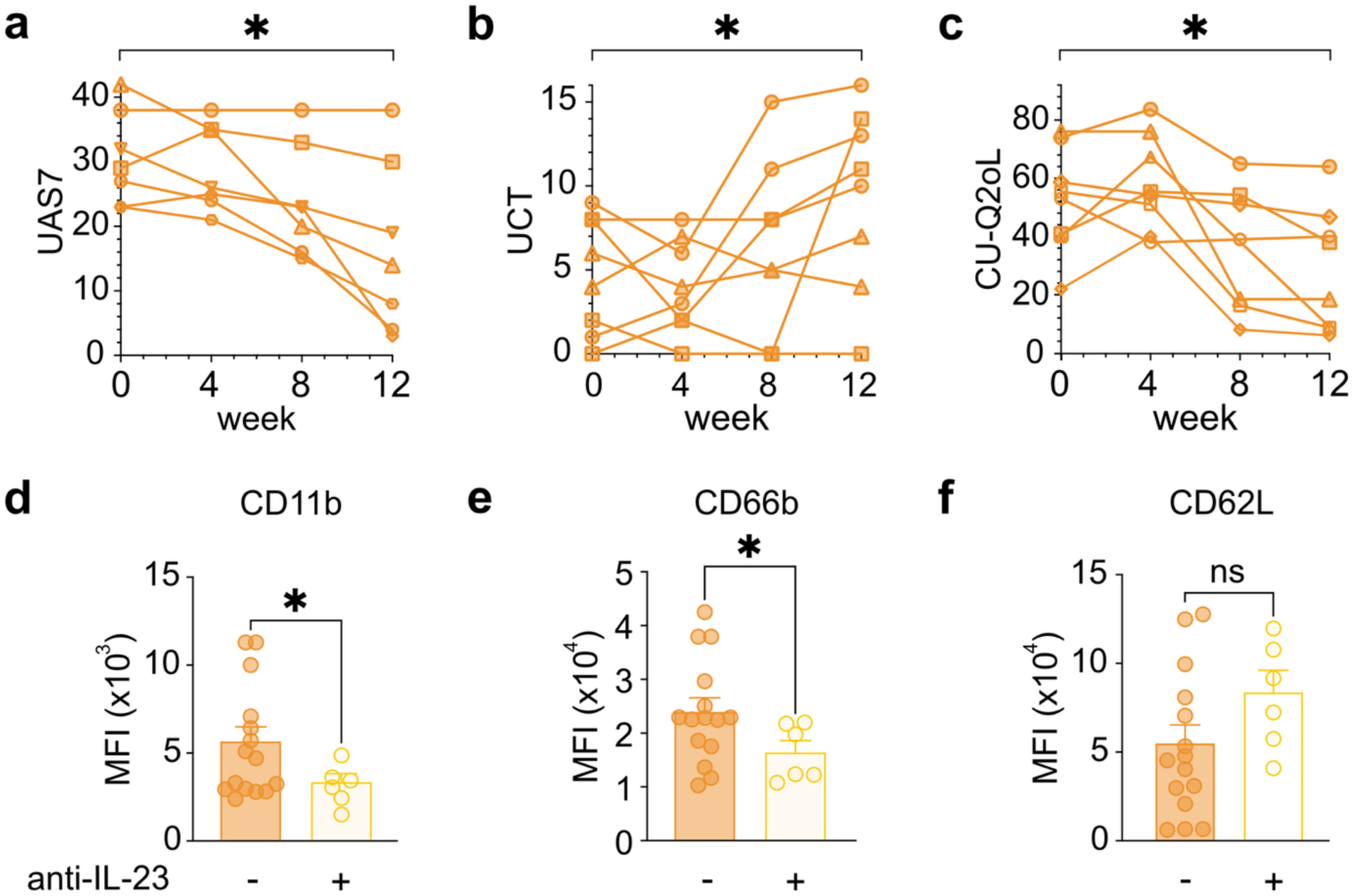
Clinical and immunological response of DPU patients treated with the anti-IL-23 antibody tildrakizumab. Therapy-refractory patients with DPU (n = 8) received 100 mg tildrakizumab subcutaneously (s.c.) at weeks 0, 4, and 12. Clinical outcomes during tildrakizumab treatment. **(a)** Urticaria Activity Score over 7 days (UAS7) (n = 7; data from one patient were not evaluable due to incomplete diary entries), **(b)** Urticaria Control Test (UCT) and **(c)** Chronic Urticaria Quality of Life Questionnaire (CU-Q2oL) at weeks 0, 4, 8 and 12. Statistical significance was assessed between week 0 and week 12. Neutrophil activation markers **(d)** CD11b, **(e)** CD66b, and **(f)** CD62L were analyzed by spectral flow cytometry in whole blood without (n = 15) tildrakizumab therapy and from DPU patients receiving tildrakizumab therapy (n = 6). Gating as described in the Figure 3.

To investigate whether these clinical findings were accompanied by differences in neutrophil activation, we compared peripheral blood neutrophils from DPU patients receiving tildrakizumab with those from DPU patients without IL-23 inhibition. CD11b and CD66b surface expression were significantly lower in the tildrakizumab-treated group (*P* = .02 and *P* = .04, respectively; Figure 5d–e). CD62L expression was higher under tildrakizumab, although this difference did not reach statistical significance (*P* = .0979; Figure 5f). This direction of change is consistent with reduced activation-associated CD62L shedding and complements the lower CD11b and CD66b expression

Together, these findings indicate that clinical improvement under IL-23 inhibition is accompanied by a less activated circulating neutrophil phenotype in therapy-refractory DPU.

### Individual Clinical Responses and Neutrophil Changes in Blood and Skin during IL-23 Inhibition

To explore how neutrophil-associated inflammation changed alongside individual clinical responses, we assessed paired peripheral blood samples from three patients before and during tildrakizumab treatment and examined changes in lesional skin. Two patients showed substantial clinical improvement, with decreased UAS7 scores, increased UCT scores, and reduced quality-of-life impairment. In contrast, the third patient showed persistently high disease activity and poor disease control, with only limited improvement in CU-Q2oL scores (Figure 6a–c).

**Figure 6:**
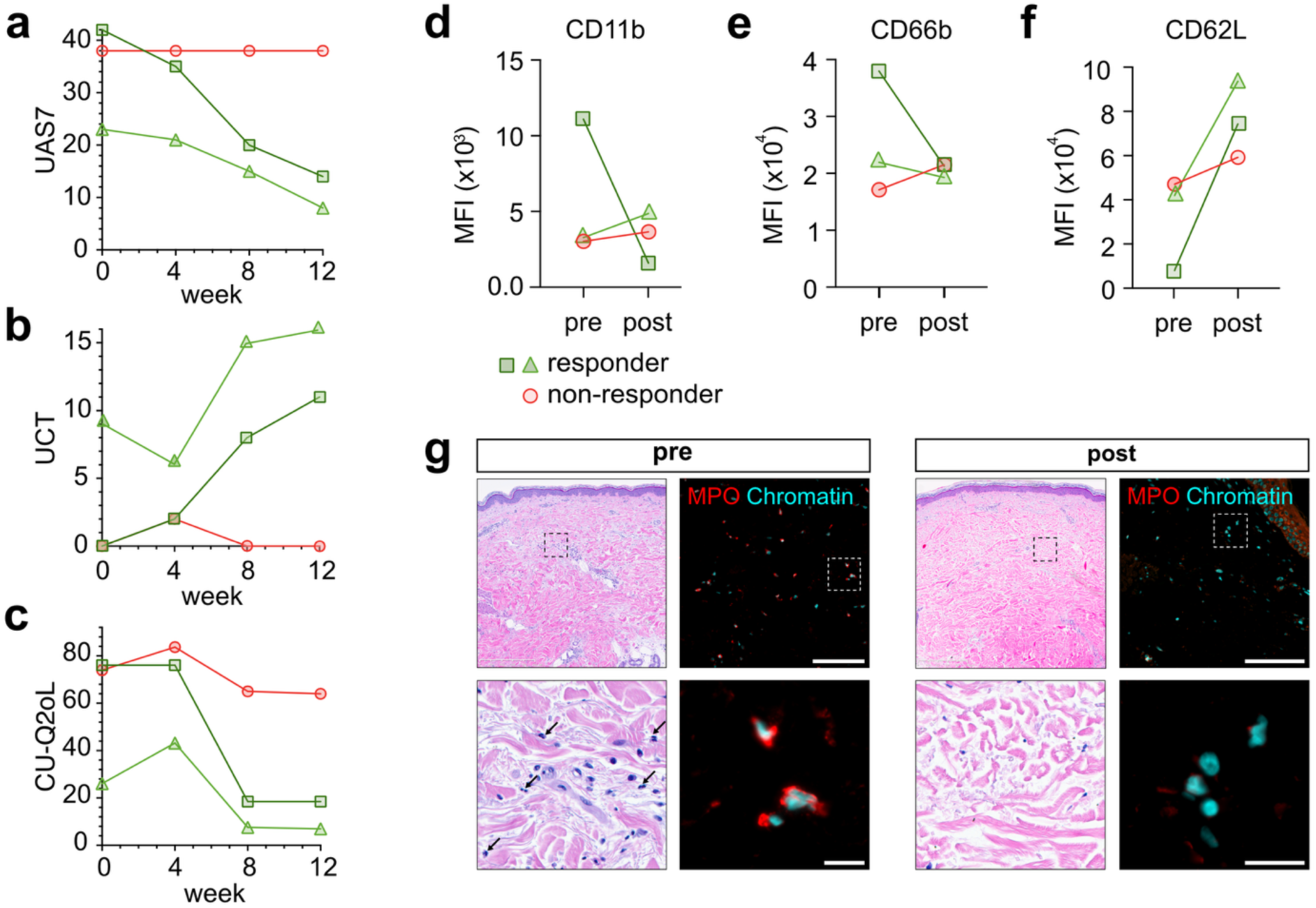
Individual paired data from DPU patients receiving tildrakizumab therapy. Clinical response was assessed using the **(a)** Urticaria Activity Score over 7 days (UAS7), (**b)** Urticaria Control Test (UCT) and **(c)** Chronic Urticaria Quality of Life Questionnaire (CU-Q2oL) scores at weeks 0, 4, 8 and 12. Each line represents one patient; responders are shown in green and the one non-responder in red. Paired pre-and post-treatment expression of **(d)** CD11b, **(e)** CD66b, and **(f)** CD62L on neutrophils from tildrakizumab-treated DPU patients (n = 3). Gating strategy as described in Figure 3. **(g)** Representative H&E staining and immunofluorescence images of lesional skin punch biopsies obtained pre-and post tildrakizumab treatment, demonstrating reduced neutrophil infiltration (representing neutrophils marked by black arrows) after treatment with tildrakizumab in one of the responders (MPO, red); chromatin is stained with DAPI (Chromatin, cyan). Scale bars Figure 6g top = 50 µm; bottom = 10 µm.

Paired flow cytometry measurements revealed changes in neutrophil activation that differed according to clinical response. CD66b expression decreased in both responders but increased in the non-responder (Figure 6f). CD11b expression decreased markedly in one responder but increased in the other responder and the non-responder (Figure 6e). CD62L expression increased in all three patients, with larger increases in the responders, consistent with reduced activation-associated shedding (Figure 6g). Among the markers examined, CD66b therefore showed the clearest correspondence with clinical response in this exploratory series.

Clinical improvement was also accompanied by a marked reduction in lesional neutrophil infiltration, illustrated by paired H&E and MPO immunofluorescence images from a clinical responder (Figure 6d). Together, these longitudinal observations complement the group-level findings by linking clinical improvement to changes in circulating neutrophil activation and reduced neutrophil infiltration in lesional skin.

## 5. DISCUSSION

In this study, we investigated the role of neutrophils in CSU and DPU by integrating analyses of lesional skin, peripheral blood, and responses to IL-23 inhibition, with the aim of identifying clinically relevant inflammatory mechanisms and potential therapeutic targets.

Our findings reveal a more pronounced neutrophil-associated inflammatory phenotype in DPU than in CSU, characterized by distinct dermal distribution patterns, enhanced NET formation, and increased peripheral neutrophil activation. The association between lesional NET formation and poorer disease control in DPU further supports the clinical relevance of these findings. Exploratory observations under IL-23 inhibition complement this picture by linking clinical improvement to changes in neutrophil activation and reduced lesional infiltration.

Together, these results support a prominent role for neutrophils in DPU and identify neutrophil-associated pathways as potential therapeutic targets in chronic urticaria.

### Deep Dermal Neutrophil Infiltration in DPU

The spatial distribution of neutrophils may help explain differences in the clinical presentation of CSU and DPU. Our findings corroborate previous histopathological observations of distinct neutrophil localization across urticaria subtypes (Haas et al., 1998) and demonstrate greater deep dermal infiltration in DPU despite comparable overall neutrophil infiltration. This distribution closely parallels the characteristic delayed, deep-seated swellings of DPU, which may persist for up to 72 hours, compared with the transient superficial wheals of CSU. The anatomical correspondence between neutrophil accumulation and the clinical phenotype suggests that tissue localization is biologically relevant rather than merely an incidental morphological finding. Together with the enhanced NET formation observed in DPU, these findings support the possibility that both the distribution and effector activity of neutrophils contribute to its prolonged inflammatory lesions.

### Pro-inflammatory Role of Neutrophils and NET Formation

We demonstrate for the first time that neutrophils in chronic urticaria undergo NET formation, thereby extending previous histopathological observations beyond the descriptive level. This finding is particularly relevant in light of previous work on Schnitzler syndrome, which reported substantial lesional NET formation but detected no NETs in the CSU comparison group (Bonnekoh et al., 2019). Our findings therefore establish NET formation as a previously unrecognized feature of chronic urticaria. Differences in patient populations, lesion characteristics, and detection methods may contribute to these contrasting observations. In our study, additional NE staining confirmed the neutrophil origin of the extracellular chromatin structures.

NETs are increasingly recognized as active amplifiers of inflammation. By exposing extracellular chromatin decorated with histones and neutrophil-derived proteases, they promote immune cell activation and establish self-perpetuating inflammatory circuits that contribute to chronic tissue inflammation (Nakabo et al., 2022; Savchenko et al., 2014). The detection of NET formation in chronic urticaria therefore suggests that neutrophilic inflammation extends beyond a transient cellular infiltrate and may actively contribute to the maintenance of local inflammation. This concept is further supported by the greater extent of NET formation in DPU and its inverse correlation with disease control (Figure 2e). Although no significant association with disease control was detected in CSU, NETs were present in both subtypes, suggesting that their contribution may depend on the inflammatory context. Together, these findings support a role for NET formation as an amplifier of urticarial inflammation, with particular relevance to DPU.

### The IL-23/IL-17 Axis as a Pathogenic Driver

The IL-23/IL-17 axis is a central inflammatory pathway in psoriasis and other inflammatory skin diseases, where IL-17A has traditionally been associated predominantly with Th17 cells. However, accumulating evidence indicates that innate immune cells, including neutrophils, represent an important source of IL-17A in psoriatic skin and may link IL-17 signalling to NET formation (Herster et al., 2020; Keijsers et al., 2014). Reports of elevated circulating IL-23 and IL-17 concentrations in CSU further implicate this pathway in urticaria (Atwa et al., 2014; Sharma et al., 2020). How this cytokine milieu relates to neutrophil-associated inflammation within urticarial lesions, however, remains poorly understood.

Our identification of IL-17A-positive neutrophils in both CSU and DPU extends the evidence for involvement of this pathway to the neutrophil compartment. No IL-17A-positive neutrophils were detected in HC skin, consistent with the absence of a neutrophilic infiltrate. IL-23-positive myeloid cells were also identified in lesional skin, in close spatial proximity to IL-17A-positive neutrophils. Although spatial proximity does not establish a direct functional interaction, this arrangement supports the possibility that locally produced IL-23 promotes neutrophil-associated IL-17 responses within the inflamed dermis. Experimental support for this mechanism comes from studies showing that IL-23 induces a distinct inflammatory phenotype in murine neutrophils, characterized by the production of IL-17A, IL-17F, and IL-22 (Li et al., 2018).

The proportion of IL-17A-positive neutrophils was similar in CSU and DPU despite their distinct dermal distribution. IL-23 positivity, expressed as the percentage of total cells, likewise showed no significant difference between subtypes, either overall or within individual dermal layers. Thus, differences in cytokine positivity alone do not explain the greater neutrophil activation and NET formation observed in DPU. However, similar proportions of cytokine-positive cells do not imply equivalent cytokine production or signalling activity, nor do they exclude a functional contribution of this pathway to either subtype. The clinical improvement and reduced neutrophil infiltration observed under IL-23 inhibition are consistent with such a contribution and suggest that a shared cytokine pathway may support different patterns or degrees of neutrophil-associated inflammation.

Instead, differences in the initially activated resident or infiltrating cells and their recruitment signals could account for the preferential accumulation of neutrophils in different dermal compartments. The more activated circulating neutrophil phenotype in DPU additionally raises the possibility of greater systemic priming, potentially enhancing effector responses after recruitment. These mechanisms may coexist and could contribute to a more prominent role for neutrophils in DPU without requiring fundamentally distinct downstream inflammatory pathways.

Moreover, despite efforts to obtain biopsies within the well-defined interval of 5 hours after lesion onset, neutrophil recruitment and activation may change over relatively short periods, and samples may capture different stages of inflammation in CSU and DPU. Time-resolved analyses could help clarify how these dynamics contribute to the distinct inflammatory patterns. Lastly, similar proportions of cytokine-positive cells do not necessarily translate into comparable cytokine production per cell or equivalent downstream signalling. Differences in cytokine abundance or availability as well as cellular responsiveness could therefore contribute to the distinct neutrophil responses even in the absence of differences in IL-17A or IL-23 positivity.

### Exploratory IL-23 Inhibition in Therapy-Refractory DPU

Building on these findings, we explored whether IL-23 inhibition was associated with biological and clinical changes in therapy-refractory DPU. The rationale for targeting IL-23 extended beyond its upstream position within the IL-23/IL-17 signalling cascade to its capacity to regulate inflammatory neutrophil responses. As discussed above, experimental studies demonstrate that IL-23 can promote a cytokine-producing neutrophil phenotype, providing a rationale for targeting the signals supporting this phenotype rather than neutralizing IL-17A alone (Li et al., 2018). Neutrophil–T-cell crosstalk may further amplify type 17 inflammation, suggesting that modulation of neutrophil-associated pathways could influence a broader inflammatory network (Macleod et al., 2023). Although its effects are not restricted to neutrophils, IL-23 inhibition therefore offers the potential to modulate several interconnected inflammatory responses.

Treatment with the IL-23p19-specific antibody tildrakizumab was associated with significant improvements in disease activity, disease control, and quality of life between baseline and week 12 (n = 8). These clinical findings were complemented by lower CD11b and CD66b expression on circulating neutrophils in patients receiving tildrakizumab compared with those without IL-23 inhibition. CD11b, the α-subunit of the integrin Mac-1, mediates neutrophil adhesion and migration and is mobilized to the cell surface upon activation. CD66b, also known as CEACAM8, is a glycoprotein located in specific granule membranes whose surface expression increases during granule exocytosis. Lower surface expression of both markers is therefore consistent with reduced neutrophil activation and granule mobilization (Connelly et al., 2022). CD62L expression was higher under treatment, consistent with reduced activation-associated shedding, although this difference did not reach statistical significance. Together, these complementary marker profiles support a less activated circulating neutrophil phenotype under IL-23 inhibition, alongside clinical improvement.

The individual longitudinal observations further connect clinical response with changes in neutrophil-associated inflammation. In three patients with paired blood samples, CD66b expression decreased in both clinical responders but increased in the non-responder, providing a biological correlate of the divergent clinical courses (Figure 6a–c, e). CD11b responses were more heterogeneous, while CD62L increased in all three patients, with larger increases in the responders (Figure 6d, f). Clinical improvement was also accompanied by a marked reduction in lesional neutrophil infiltration, illustrated by paired biopsies from a responder (Figure 6g). Together, these observations in blood and skin strengthen the biological relevance of the neutrophil changes associated with treatment. Whether these changes reflect direct modulation of neutrophils or broader effects of IL-23 inhibition on the inflammatory milieu remains to be determined. The correspondence between CD66b changes and clinical response also supports prospective evaluation of circulating neutrophil profiles for treatment monitoring and investigation of their potential to inform individualized therapy across different CU inflammatory phenotypes.

Our findings extend previous observations of clinical improvement under tildrakizumab in therapy-refractory CSU to DPU and complement them with measurements of neutrophil activation and tissue infiltration (Bonnekoh et al., 2023). They raise the possibility that targeting IL-23 may address an inflammatory component of DPU that remains insufficiently controlled by existing treatment approaches.

The small observational case series and absence of a clinical control group preclude firm conclusions regarding treatment efficacy. The lower activation-marker expression in the treated group cannot by itself establish a treatment-induced change; the paired measurements provide longitudinal evidence in three patients. The biological changes observed under IL-23 inhibition may involve both direct effects on neutrophils and modulation of the surrounding inflammatory network; their respective contributions warrant further investigation.

Further investigation of NET formation during treatment could extend our understanding of how IL-23 inhibition modulates neutrophil effector responses. Our findings already provide the first exploratory evidence linking IL-23 inhibition to clinical improvement and changes in neutrophil-associated inflammation in therapy-refractory DPU, supporting further investigation of this pathway in controlled clinical studies.

### Conclusions and Future Perspectives

Our findings also have implications for disease monitoring and therapeutic stratification. Current approaches to CSU endotyping incorporate clinical assessments and laboratory parameters such as total IgE and anti-TPO antibodies, which are associated with treatment response but do not consistently predict individual outcomes (Fok et al., 2021; Kolkhir et al., 2021; Wong et al., 2025). The continuing need for reliable biomarkers provides a rationale for investigating additional inflammatory profiles that could inform individualized treatment (Gimenez-Arnau et al., 2024; Kulumbegov et al., 2023; Ridge et al., 2025; Sabag et al., 2020). Our tissue findings and circulating neutrophil activation profiles provide candidates for prospective evaluation, both for monitoring biological responses and for investigating whether pretreatment profiles identify patients likely to benefit from targeted intervention (Prosty et al., 2022; Sabag et al., 2020).

Taken together, this study identifies neutrophils as active contributors to inflammation in chronic urticaria, with particularly pronounced tissue and circulating activation profiles in DPU. Distinct dermal distribution patterns and enhanced NET formation support a more prominent contribution of neutrophils to DPU than to CSU. The identification of IL-17A-positive neutrophils and IL-23-positive myeloid cells further implicates this pathway in urticarial inflammation. Exploratory observations provide the first in vivo evidence linking IL-23 inhibition to clinical improvement and changes in neutrophil-associated inflammation in therapy-refractory DPU. Together, these findings support further evaluation of neutrophil-associated pathways as therapeutic targets and provide a foundation for more individualized treatment approaches in chronic urticaria.

## Data Availability

All data produced in the present study are available upon reasonable request to the authors

## ACKNOWLEDGMENETS

We thank all patients and healthy donors who made this study possible. LE was supported by the Deutsche Forschungsgemeinschaft (DFG, German Research Foundation) – Project-ID 431460824 – CRC 1450. MB was supported by the Alfred-Marchionini Foundation and the Deutsche Dermatologische Gesellschaft (DDG). The project also received seed funding from Almirall. Almirall had no role in study design, data collection, analysis or interpretation, manuscript preparation, or the decision to submit the manuscript for publication. We thank Kerstin Steinbrink, Undine Lippert, Stefan Frischbutter, Verena Lorenz and Wolfgang Pfützner for helpful input and discussions.

## List of Abbreviations

aiCSU;: Autoimmune Chronic Spontaneous Urticaria (Type IIb)
anti-TPO;: Anti-Thyroid Peroxidase Antibody
ASST;: Autologous Serum Skin Test
AU;: Acute Urticaria
BAT;: Basophil Activation Test
CindU;: Chronic Inducible Urticaria
CRP;: C-Reactive Protein
CSU;: Chronic Spontaneous Urticaria
CU;: Chronic Urticaria
CU-Q2oL;: Chronic Urticaria Quality of Life Questionnaire
DPU;: Delayed Pressure Urticaria
EDTA;: Ethylenediaminetetraacetic Acid
ELISA;: Enzyme-Linked Immunosorbent Assay
FACS;: Fluorescence-Activated Cell Sorting
H3cit;: Citrullinated Histone H3
HBSS;: Hank’s Balanced Salt Solution
HC;: Healthy Controls
IFN;: Interferon
IgE;: Immunoglobulin E
IgG;: Immunoglobulin G
IL;: Interleukin
MFI;: Mean/Median Fluorescence Intensity
MIT;: Mast Cell Intracellular Trap
MPO;: Myeloperoxidase
NET;: Neutrophil Extracellular Trap
NETosis;: Neutrophil Extracellular Trap Formation
NUD;: Neutrophilic Urticarial Dermatosis
RBC;: Red Blood Cell
ROI;: Region of Interest
SEM;: Standard Error of the Mean
TNF-α;: Tumor Necrosis Factor Alpha
UAS7;: Urticaria Activity Score over 7 days
UCT;: Urticaria Control Test

## Notes

### Competing Interest Statement

The authors have declared no competing interest.

### Author Declarations

ethical approval by the Landesärztekammer Westfalen-Lippe (reference 2022-420-f-S).

## REFERENCES

1. Bernstein, J. A., Lang, D. M., Khan, D. A., Craig, T., Dreyfus, D., Hsieh, F., Sheikh, J., Weldon, D., Zuraw, B., Bernstein, D. I., Blessing-Moore, J., Cox, L., Nicklas, R. A., Oppenheimer, J., Portnoy, J. M., Randolph, C. R., Schuller, D. E., Spector, S. L., Tilles, S. A., & Wallace, D. (2014). The diagnosis and management of acute and chronic urticaria: 2014 update. The Journal of allergy and clinical immunology, 133(5), 1270–1277.e1266. 10.1016/J.JACI.2014.02.036

2. Bonnekoh, H., Scheffel, J., Wu, J., Hoffmann, S., Maurer, M., & Krause, K. (2019). Skin and systemic inflammation in Schnitzler’s syndrome are associated with neutrophil extracellular trap formation. Frontiers in Immunology, 10(MAR), 546–546. 10.3389/fimmu.2019.00546

3. Dudeck, J., Kotrba, J., Immler, R., Hoffmann, A., Voss, M., Alexaki, V. I., Morton, L., Jahn, S. R., Katsoulis-Dimitriou, K., Winzer, S., Kollias, G., Fischer, T., Nedospasov, S. A., Dunay, I. R., Chavakis, T., Müller, A. J., Schraven, B., Sperandio, M., & Dudeck, A. (2021). Directional mast cell degranulation of tumor necrosis factor into blood vessels primes neutrophil extravasation. Immunity, 54(3), 468–483.e465. 10.1016/J.IMMUNI.2020.12.017

4. Erpenbeck, L., Chowdhury, C., Zsengellér, Z. K., Gallant, M., Burke, S. D., Cifuni, S., Hahn, S., Wagner, D. D., & Karumanchi, S. A. (2016). PAD4 Deficiency Decreases Inflammation and Susceptibility to Pregnancy Loss in a Mouse Model. Biology of reproduction, 95(6), 1–11. 10.1095/BIOLREPROD.116.140293

5. Erpenbeck, L., Gruhn, A. L., Kudryasheva, G., Günay, G., Meyer, D., Busse, J., Neubert, E., Schön, M. P., Rehfeldt, F., & Kruss, S. (2019). Effect of Adhesion and Substrate Elasticity on Neutrophil Extracellular Trap Formation. Frontiers in Immunology, 10. 10.3389/FIMMU.2019.02320

6. Erpenbeck, L., & Schön, M. P. (2017). Neutrophil extracellular traps: Protagonists of cancer progression? In Oncogene (Vol. 36, pp. 2483–2490): Nature Publishing Group.

7. Fok, J. S., Kolkhir, P., Church, M. K., Maurer, M., Correspondence, M., Maurer, D., & Allergology, A.-C.-C. (2021). Predictors of treatment response in chronic spontaneous urticaria. 10.1111/all.14757

8. Fuchs, T. A., Abed, U., Goosmann, C., Hurwitz, R., Schulze, I., Wahn, V., Weinrauch, Y., Brinkmann, V., & Zychlinsky, A. (2007). Novel cell death program leads to neutrophil extracellular traps. The Journal of cell biology, 176(2), 231–241. 10.1083/JCB.200606027

9. Giménez-Arnau, A. M., DeMontojoye, L., Asero, R., Cugno, M., Kulthanan, K., Yanase, Y., Hide, M., & Kaplan, A. P. (2021). The Pathogenesis of Chronic Spontaneous Urticaria: The Role of Infiltrating Cells. The Journal of Allergy and Clinical Immunology: In Practice, 9(6), 2195–2208. 10.1016/J.JAIP.2021.03.033

10. Gimenez-Arnau, A. M., Salman, A., & Podder, I. (2024). Biomarkers to predict therapeutic response in chronic spontaneous urticaria: a review. European journal of dermatology : EJD, 34(1), 3–12. 10.1684/EJD.2024.4600

11. Gonçalo, M., Gimenéz-Arnau, A., Al-Ahmad, M., Ben-Shoshan, M., Bernstein, J. A., Ensina, L. F., Fomina, D., Galvàn, C. A., Godse, K., Grattan, C., Hide, M., Katelaris, C. H., Khoshkhui, M., Kocatürk, E., Kulthanan, K., Medina, I., Nasr, I., Peter, J., Staubach, P.,…Maurer, M. (2021). The global burden of chronic urticaria for the patient and society. British Journal of Dermatology, 184(2), 226–236. 10.1111/BJD.19561

12. Grundmann, S. A., Kiefer, S., Luger, T. A., & Brehler, R. (2011). Delayed pressure urticaria -Dapsone heading for first-line therapy? JDDG -Journal of the German Society of Dermatology, 9(11), 908–912. 10.1111/J.1610-0387.2011.07749.X

13. Haas, N., Toppe, E., & Henz, B. M. (1998). Microscopic Morphology of Different Types of Urticaria. Archives of Dermatology, 134(1), 41–46. 10.1001/ARCHDERM.134.1.41

14. Herster, F., Bittner, Z., Archer, N. K., Dickhöfer, S., Eisel, D., Eigenbrod, T., Knorpp, T., Schneiderhan-Marra, N., Löffler, M. W., Kalbacher, H., Vierbuchen, T., Heine, H., Miller, L. S., Hartl, D., Freund, L., Schäkel, K., Heister, M., Ghoreschi, K., & Weber, A. N. R. (2020). Neutrophil extracellular trap-associated RNA and LL37 enable self-amplifying inflammation in psoriasis. Nature Communications, 11(1). 10.1038/S41467-019-13756-4

15. Hollstein, M. M., Manzke, V., Scheidmann, S. E. F., Schrenker, S., Schaffrinski, M., Neubert, E., Kramer, D., Raker, V. K., Schön, M. P., & Erpenbeck, L. (2023). Targeting neutrophil extracellular traps (NETs) ameliorates inflammation in murine psoriasiform dermatitis. Journal of Dermatological Science, 109(3), 143–146. 10.1016/J.JDERMSCI.2023.02.006

16. Kahlenberg, J. M., Carmona-Rivera, C., Smith, C. K., & Kaplan, M. J. (2013). Neutrophil extracellular trap-associated protein activation of the NLRP3 inflammasome is enhanced in lupus macrophages. Journal of Immunology (Baltimore, Md. : 1950), 190(3), 1217–1226. 10.4049/JIMMUNOL.1202388

17. Keijsers, R. R. M. C., Hendriks, A. G. M., Van Erp, P. E. J., Van Cranenbroek, B., Van De Kerkhof, P. C. M., Koenen, H. J. P. M., & Joosten, I. (2014). In vivo induction of cutaneous inflammation results in the accumulation of extracellular trap-forming neutrophils expressing RORγt and IL-17. Journal of Investigative Dermatology, 134(5), 1276–1284. 10.1038/jid.2013.526

18. Kulumbegov, B., Chikovani, T., Gotua, M., Kikodze, N., & Magen, E. (2023). Interleukin-33, endothelin-1, and inflammatory parameters in chronic spontaneous urticaria. Allergy and Asthma Proceedings, 44(6), 429–435. 10.2500/AAP.2023.44.230051

19. Lee, S. J., Ha, E. K., Jee, H. M., Lee, K. S., Lee, S. W., Kim, M. A., Kim, D. H., Jung, Y. H., Sheen, Y. H., Sung, M. S., & Han, M. Y. (2017). Prevalence and Risk Factors of Urticaria With a Focus on Chronic Urticaria in Children. *Allergy*, Asthma & Immunology Research, 9(3), 212–212. 10.4168/AAIR.2017.9.3.212

20. Martinod, K., Witsch, T., Erpenbeck, L., Savchenko, A., Hayashi, H., Cherpokova, D., Gallant, M., Mauler, M., Cifuni, S. M., & Wagner, D. D. (2017). Peptidylarginine deiminase 4 promotes age-related organ fibrosis. The Journal of Experimental Medicine, 214(2), 439–458. 10.1084/JEM.20160530

21. Maurer, M., Abuzakouk, M., Bérard, F., Canonica, W., Oude Elberink, H., Giménez-Arnau, A., Grattan, C., Hollis, K., Knulst, A., Lacour, J. P., Lynde, C., Marsland, A., McBride, D., Nakonechna, A., Ortiz de Frutos, J., Proctor, C., Sussman, G., Sweeney, C., Tian, H.,…Balp, M. M. (2017). The burden of chronic spontaneous urticaria is substantial: Real-world evidence from ASSURE-CSU. Allergy, 72(12), 2005–2016. 10.1111/ALL.13209

22. Mihlan, M., Wissmann, S., Gavrilov, A., Kaltenbach, L., Britz, M., Franke, K., Hummel, B., Imle, A., Suzuki, R., Stecher, M., Glaser, K. M., Lorentz, A., Carmeliet, P., Yokomizo, T., Hilgendorf, I., Sawarkar, R., Diz-Muñ, A., Buescher, J. M., Mittler, G.,…Lämmermann, T. (2024). Neutrophil trapping and nexocytosis, mast cell-mediated processes for inflammatory signal relay. Cell, 187(19), 5316–5335.e5328. 10.1016/j.cell.2024.07.014

23. Morioke, S., Takahagi, S., Iwamoto, K., Shindo, H., Mihara, S., Kameyoshi, Y., & Hide, M. (2010). Pressure challenge test and histopathological inspections for 17 Japanese cases with clinically diagnosed delayed pressure urticaria. Archives of Dermatological Research, 302(8), 613–617. 10.1007/s00403-010-1053-4

24. Nakabo, S., Romo-Tena, J., & Kaplan, M. J. (2022). Neutrophils as Drivers of Immune Dysregulation in Autoimmune Diseases with Skin Manifestations. Journal of Investigative Dermatology, 142(3), 823–833. 10.1016/J.JID.2021.04.014

25. Neubert, E., Meyer, D., Rocca, F., Günay, G., Kwaczala-Tessmann, A., Grandke, J., Senger-Sander, S., Geisler, C., Egner, A., Schön, M. P., Erpenbeck, L., & Kruss, S. (2018). Chromatin swelling drives neutrophil extracellular trap release. Nature Communications *2018 9:1*, 9(1), 3767. 10.1038/s41467-018-06263-5

26. Neubert, E., Senger-Sander, S. N., Manzke, V. S., Busse, J., Polo, E., Scheidmann, S. E. F., Schön, M. P., Kruss, S., & Erpenbeck, L. (2019). Serum and Serum Albumin Inhibit in vitro Formation of Neutrophil Extracellular Traps (NETs). Frontiers in Immunology, 10(JAN). 10.3389/FIMMU.2019.00012

27. O’Donnell, B. F., Lawlor, F., Simpson, J., Morgan, M., & Greaves, M. W. (1997). The impact of chronic urticaria on the quality of life. Br J Dermatol, 136(2), 197–201.

28. Prosty, C., Gabrielli, S., Ben-Shoshan, M., Le, M., Giménez-Arnau, A. M., Litvinov, I. V., Lefrançois, P., & Netchiporouk, E. (2022). In silico Identification of Immune Cell-Types and Pathways Involved in Chronic Spontaneous Urticaria. Frontiers in Medicine, 9. 10.3389/FMED.2022.926753/FULL

29. Ridge, K., Ahmad, R., Moran, Barry, Cliona, O. f., Dunne, J., Conor, Finlay, M., Irvine, A. D., & Conlon, N. (2025). Clinical & Experimental Allergy Towards Personalised Therapy in Chronic Spontaneous Urticaria: Advancing From Endotype to Clinical Response. Clinical & Experimental Allergy, 55, 1070–1082. 10.1111/cea.70100

30. Sabag, D. A., Matanes, L., Bejar, J., Sheffer, H., Barzilai, A., Church, M. K., Toubi, E., Maurer, M., & Vadasz, Z. (2020). Interleukin-17 is a potential player and treatment target in severe chronic spontaneous urticaria. Clinical & Experimental Allergy, 50(7), 799–804. 10.1111/CEA.13616

31. Savchenko, A. S., Borissoff, J. I., Martinod, K., De Meyer, S. F., Gallant, M., Erpenbeck, L., Brill, A., Wang, Y., & Wagner, D. D. (2014). VWF-mediated leukocyte recruitment with chromatin decondensation by PAD4 increases myocardial ischemia/reperfusion injury in mice. Blood, 123(1), 141–148. 10.1182/BLOOD-2013-07-514992

32. Schön, M. P., Broekaert, S. M. C., & Erpenbeck, L. (2017). Sexy again: the renaissance of neutrophils in psoriasis. Experimental Dermatology, 26(4), 305–311. 10.1111/EXD.13067

33. Schön, M. P., & Erpenbeck, L. (2018). The Interleukin-23/Interleukin-17 Axis Links Adaptive and Innate Immunity in Psoriasis. Frontiers in Immunology, 9(JUN), 1–1. 10.3389/FIMMU.2018.01323

34. Weller, K., Maurer, M., Bauer, A., Wedi, B., Wagner, N., Schliemann, S., Kramps, T., Baeumer, D., Multmeier, J., Hillmann, E., & Staubach, P. (2022). Epidemiology, comorbidities, and healthcare utilization of patients with chronic urticaria in Germany. Journal of the European Academy of Dermatology and Venereology, 36(1), 91–99. 10.1111/JDV.17724

35. Wienkamp, A. K., Erpenbeck, L., & Rossaint, J. (2022). Platelets in the NETworks interweaving inflammation and thrombosis. Front Immunol, 13, 953129. 10.3389/fimmu.2022.953129

36. Zuberbier, T., Abdul Latiff, A. H., Abuzakouk, M., Aquilina, S., Asero, R., Baker, D., Ballmer-Weber, B., Bangert, C., Ben-Shoshan, M., Bernstein, J. A., Bindslev-Jensen, C., Brockow, K., Brzoza, Z., Chong Neto, H. J., Church, M. K., Criado, P. R., Danilycheva, I. V., Dressler, C., Ensina, L. F.,…Maurer, M. (2022). The international EAACI/GA²LEN/EuroGuiDerm/APAAACI guideline for the definition, classification, diagnosis, and management of urticaria. Allergy, 77(3), 734–766. 10.1111/ALL.15090

37. Zuberbier, T., Ensina, L. F., Gimenez-Arnau, A., Grattan, C., Kocaturk, E., Kulthanan, K., Kolkhir, P., & Maurer, M. (2024). Chronic urticaria: unmet needs, emerging drugs, and new perspectives on personalised treatment. Lancet, 404(10450), 393–404. 10.1016/S0140-6736(24)00852-3

