## Supplementary Information for "Neutrophils Link Tissue Inflammation to Disease Control in Delayed Pressure Urticaria"

*Marie Britz[1]*, Sangeetha Shankar[1]*, Franziska Wulfert[1], Meike Steinert[1], Caroline Constanze Albers[1], Michael Wojak[1], Soobin Dokko[1], Mathis Richter[2], Michael P. Schön[3,4], Mathias Sulk[1#], Luise Erpenbeck[1#]*

[1]Department of Dermatology, University Hospital Münster, Münster, Germany
[2]Institute for Experimental Pathology (ExPat), ZMBE, University of Münster, Münster, Germany
[3] Department of Dermatology, Venereology and Allergology, University Medical Center Göttingen, Göttingen, Germany
[4] Lower Saxony Institute of Occupational Dermatology, University Medical Center Göttingen, Göttingen, Germany
* Shared first-authorship, # Shared last-authorship

S1

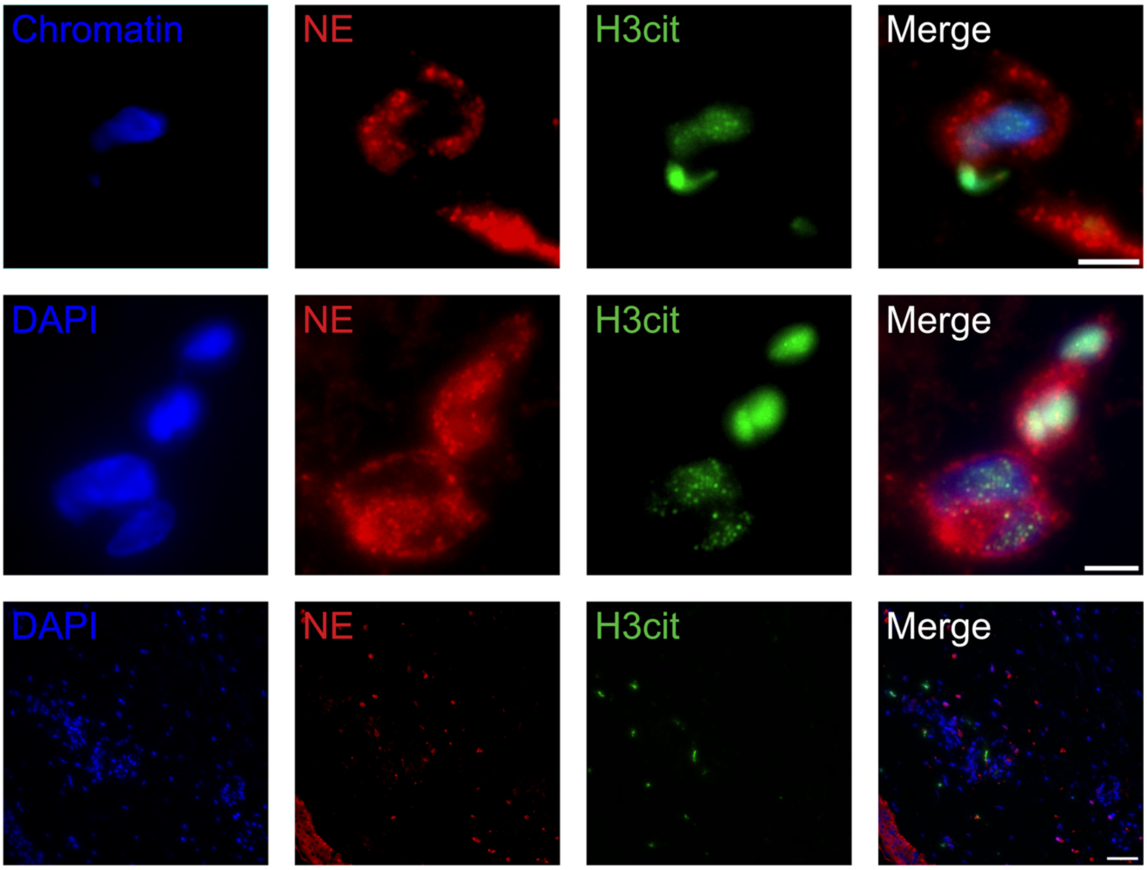


***Supplementary Figure 1: Immunofluorescence staining of neutrophil extracellular traps (NETs) in a representative DPU skin biopsy.*** *Sections were stained with fluorescently labelled antibodies against neutrophil elastase (NE; red) and H3cit (green); nuclei were counterstained with DAPI (blue). Colocalization of NE, H3cit and DNA corroborates the identity of extracellular traps released by neutrophils. Scale bars upper two rows: 5 µm. Lower row: 50µm.*

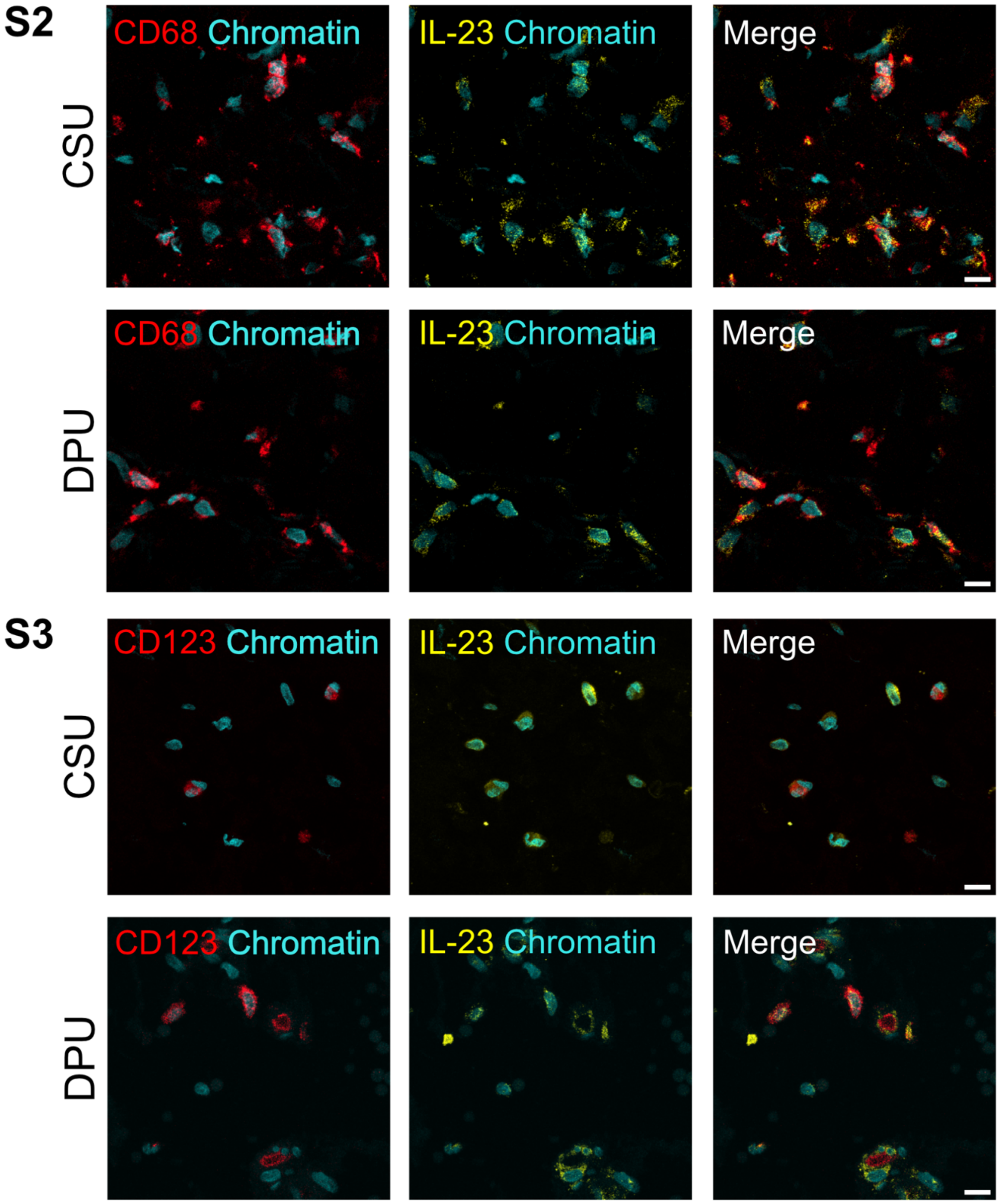


***Supplementary Figure 2****: Representative immunofluorescence images of CD68- and CD123 (both in red)-positive cells expressing IL-23 (yellow) in lesional DPU skin. The scale bar represents XXX µm.*
